# Carrageenan-containing nasal spray alleviates allergic symptoms in subjects with grass pollen allergy: A randomized, controlled, crossover clinical trial

**DOI:** 10.1101/2023.10.19.23297206

**Authors:** Nicole Unger-Manhart, Martina Morokutti-Kurz, Petra Zieglmayer, Patrick Lemell, Markus Savli, René Zieglmayer, Eva Prieschl-Grassauer

## Abstract

**Purpose:** Nonpharmacological nasal sprays forming a barrier between allergens and the nasal mucosa are used to manage symptoms of allergic rhinitis. We aimed to evaluate the safety and effectiveness of Callergin, a nasal spray containing barrier-forming iota-carrageenan, in the treatment of allergic rhinitis.

**Methods:** In this randomized, controlled, crossover trial, we assigned adults with grass pollen allergy to receive Callergin, VisAlpin and no treatment in a random order for three consecutive periods, separated by a washout period of 7 days. Subjects prophylactically applied one puff of nasal spray to each nostril 5-10 minutes prior to challenge. The primary endpoint was mean change from baseline in ‘total nasal symptom score’ (TNSS) over 3 hours, a sum of rhinorrhea, itching, sneezing, and congestion scores, recorded every 15 minutes during the challenge period.

**Results:** A total of 42 subjects underwent randomization. Exposure to grass pollen for 3 hours led to a notable TNSS increase from baseline in all subjects at all times. Mean TNSS was lower when subjects received treatment with Callergin compared to no treatment, although the difference did not reach statistical significance (untreated 6.96 ± 2.30; Callergin 6.59 ± 1.93; difference 0.37 points [95% CI -0.17 to 0.91]; p=0.170). In a post-hoc analysis, mean TNSS at 3 hours was significantly reduced with Callergin treatment compared to no treatment (untreated 8.29 ± 2.64; Callergin 7.70 ± 2.56; difference 0.60 points [95% CI -0.10 to 1.29] p=0.028). While all individual nasal symptoms contributed to this effect, rhinorrhea (p=0.013) and congestion (p=0.076) contributed the most. Consistently, nasal secretion weight was slightly reduced with Callergin (p=0.119). VisAlpin improved nasal symptoms, but not significantly in either analysis.

The incidence of adverse events was similar among treatment groups.

**Conclusion:** Prophylactic treatment with Callergin is safe and alleviates nasal symptoms in adults with grass pollen allergy.

**Trial registration:** NCT04531358

## Introduction

Allergic rhinitis (AR) is an immunoglobulin E (IgE)-mediated hypersensitivity reaction occurring after exposure to airborne allergens.^1,2^ The classic symptoms are nasal itching, obstruction, sneezing and rhinorrhea (runny nose), although some subjects may also experience ocular or upper respiratory symptoms. ^3,4^

About half of allergic subjects have AR symptoms for more than four months/year and one-fifth for more than nine months/year. If not treated properly, these symptoms affect people’s quality of life and are associated with substantial healthcare costs (eg, exacerbations of sinusitis and asthma, nasal polyps, hearing impairment) and other economic impacts (eg, less productivity).^5^ Current treatment recommendations include pharmacotherapy with oral or intranasal H_1_-antihistamines, intranasal corticosteroids, or a combination of intranasal H_1_-antihistamines and corticosteroids, as well as allergen avoidance and immunotherapy.

Drug-free, locally acting, barrier-forming nasal sprays can be an attractive alternative treatment for mild to moderate allergic rhinitis and in special populations (eg, pregnant women or children), or as a complementary therapy for moderate to severe allergic rhinitis. Callergin is a nasal spray based on iota-carrageenan (Carragelose^®^), a natural polymer from red seaweed, which lines the nasal mucosa to prevent contact with airborne allergens. ^6,7^ Carragelose^®^ is certified for marketing in the EU, parts of Asia and Australia, as a component of nasal sprays, throat sprays and lozenges. Callergin nasal spray is classified as a class I substance-based medical device.

The objective of this study was to evaluate the clinical performance of Callergin nasal spray in preventing AR symptoms in subjects with grass pollen allergy when compared to a “no-treatment” control. VisAlpin Alpensalz, a marketed saline nasal spray, was added as a comparator in this study because it can be used as an accompanying treatment for stuffy nose caused by a cold or allergy. ^8^

## Methods

### Study design

This was a randomized, open-label no-treatment-controlled, double-blind active-controlled, three-period crossover trial to assess the safety and efficacy of Callergin nasal spray in subjects with AR caused by grass pollen (NCT04531358). In each treatment period, participants received either Callergin, the comparator VisAlpin Alpensalz, or no treatment at all, separated by a washout period of 7 days. The two treatment groups were double blinded against each other, while the untreated group was naturally unblinded. The study was performed at the Vienna Challenge Chamber in Vienna, Austria. The Ethics Committee City of Vienna oversaw trial conduct and documentation.

### Participants

Eligible patients were aged 18-65 and had a documented history of moderate-to-severe seasonal AR to grass pollen with or without mild-to-moderate asthma. At screening, participants had to score ≥6 in total nasal symptom score (TNSS) in response to approximately 1500 grass pollen grains/m^3^ within the first 2 hours inside the exposure chamber. Main exclusion criteria comprised: uncontrolled asthma within the past 3 months; current upper respiratory tract infection, sinusitis or otitis media; presence of clinically relevant nasal polyps; history of tuberculosis; previous or ongoing immunotherapy to grass pollen. Treatment with steroids, long-lasting antihistamines, leukotriene antagonists, mast cell stabilizers, or nasal decongestants was not permitted.

### Randomization and masking

Patients were randomly assigned to one of the three treatments per treatment period using a crossover randomization with balanced blocks. Double blinding was guaranteed by identical presentation of the nasal sprays and the use of neutral randomization numbers for the differentiation of the packs.

### Interventions and procedures

During each treatment period, participants administered one puff (140 μl) of either Callergin or the comparator VisAlpin Alpensalz in each nostril 5-10 minutes before entering an environmental exposure chamber or remained untreated for that period. Study drug administration was supervised by study staff and recorded in the accountability log. Participants were then exposed to a standard grass pollen allergen mixture (1500 grass pollen grains/m^3^, Allergopharma) for 3 hours.

### Endpoints

The primary efficacy endpoint was mean change from baseline in ‘total nasal symptom score’ (TNSS) over 3 hours. TNSS is the sum of the nasal symptoms of congestion, rhinorrhea, itching and sneezing. Each symptom was rated on a scale from 0 to 3, whereas “0” corresponded to “no symptoms”, “1” to “mild symptoms” (easy to tolerate), “2” to “moderate symptoms” (bothersome, but tolerable) and “3” to “severe symptoms” (hard to tolerate). Secondary endpoints included change from baseline in ‘total ocular symptom score’ (TOSS; eye symptoms: itchy eyes, red eyes and watery eyes) and ‘total respiratory symptom score’ (TRSS; respiratory symptoms: cough, wheeze and dyspnea). During their 3-hour stay in the challenge chamber, participants were asked to rate their nasal, ocular, and respiratory allergy symptoms at 15-minute intervals. Nasal secretion was evaluated by weighing paper tissues used by the subjects during their stay in the chamber and collected every 30 minutes. Nasal airflow was measured by rhinomanometry at 30, 60, 120 and 180 minutes. Safety assessments included measurement of vital signs and lung function (hourly measurement of FEV1 by spirometry) during treatment visits, as well as electrocardiogram, physical examination, nasal examination and blood analysis at screening and follow-up visits. All safety analyses were based on the safety population defined as all subjects starting the challenge provocation qualification session.

### Statistical analysis

Sample size was calculated to reach apower of 90% with an alpha level p≤0.05 resulting in 36 subjects needed for evaluation. Considering a dropout rate of 10-15%, 50 subjects had to be screened to randomize about 42 subjects to get at least 36 evaluable subjects at the end of the trial. For the primary efficacy variable, the within participant comparison of Callergin versus no treatment was performed using a three-period analysis of variance (ANOVA) appropriate for the crossover design. Period was included in the analysis model as a fixed effect to confirm the assumption of no period effect. Subject was included in the model as a random effect. A 95% confidence interval was calculated for the difference in means between the two active treatments from a two-sided paired t-test. The Tukey procedure was applied for post-hoc comparisons to adjust for multiplicity. The hypothesis to be tested was superiority of Callergin spray in comparison to the no-treatment control. The null hypothesis is defined as

- [mean TNSS (Test)] ≥ [mean TNSS (untreated condition)]

The alternative hypothesis was defined as

- [mean TNSS (Test)] < [mean TNSS (untreated condition)]

The effects of VisAlpin spray were only described in an explorative manner. Therefore, superiority/non-inferiority of Callergin vs. VisAlpin was not defined for this study.

## Results

Between September 2020 and December 2020, a total of 47 subjects with grass pollen allergy were screened after giving informed consent. Of these, 42 participants were randomized to six possible treatment sequences, with each participant acting as her/his own control. All randomized participants completed the study and were included in the analysis (Figure 1).

**Figure 1:**
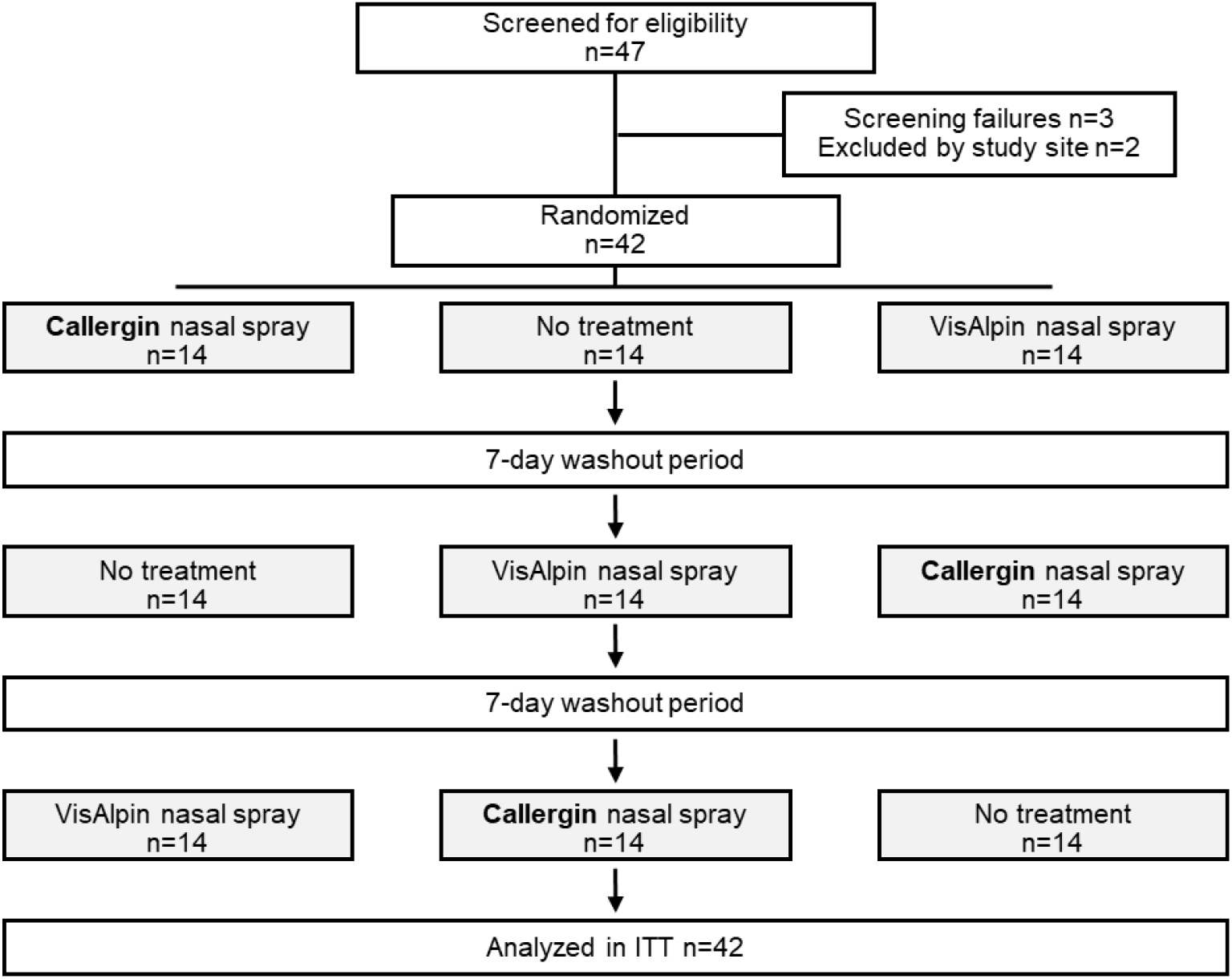
Patient disposition: Among the 5 patients who were not randomized, 3 failed to respond to the allergen challenge on screening and 2 were lost to follow-up. Randomized participants were assigned to one of six possible treatment sequences.

Baseline characteristics were balanced across the six treatment sequences (data not shown) After each allergen challenge, TNSS scores had returned to their baseline values at the start of the following treatment period, excluding a potential carry-over effect from one treatment period into the subsequent one (ANOVA model, p-value>0.05) (Table 1).

**Table 1:** Demographic and clinical characteristic of the patients at baseline (intention-to-treat population) Abbreviations: BMI, body mass index, TNSS, total nasal symptom score.

|  | All patients<br>(n=42) |
| --- | --- |
| Sex: |  |
| Male | 16 (38%) |
| Female | 26 (62%) |
| Ethnicity: |  |
| Caucasian | 42 (100%) |
| Age (years) | 36 (22-62) |
| BMI (kg/m<sup>2</sup>) | 24 (19-30) |
| Grass pollen allergy: |  |
| TNSS score in treatment period 1 - at baseline | 0.14 ± 0.42 |
| TNSS score in treatment period 2 - at baseline | 0.12 ± 0.40 |
| TNSS score in treatment period 3 - at baseline | 0.12 ± 0.40 |

All subjects suffered from increasing nasal symptoms (mean change from baseline in TNSS: untreated 6.96 ± 2.30; Callergin 6.59 ± 1.93; VisAlpin 6.34 ± 1.77) and a 40-45% reduction in nasal airflow during the three hours they were exposed to grass pollen. In contrast, too few subjects developed significant eye and respiratory symptoms to allow statistical evaluation. The increase in TNSS following allergen exposure over 3 hours was slightly lower with Callergin treatment than without treatment (Δ=0.37, 95% CI -0.17 to 0.91; p=0.17). Our results obtained with the comparator VisAlpin showed a similar trend but did not reach statistical significance either (Δ=0.61, 95% CI -0.17 to 0.91; Figure 2).

**Figure 2:**
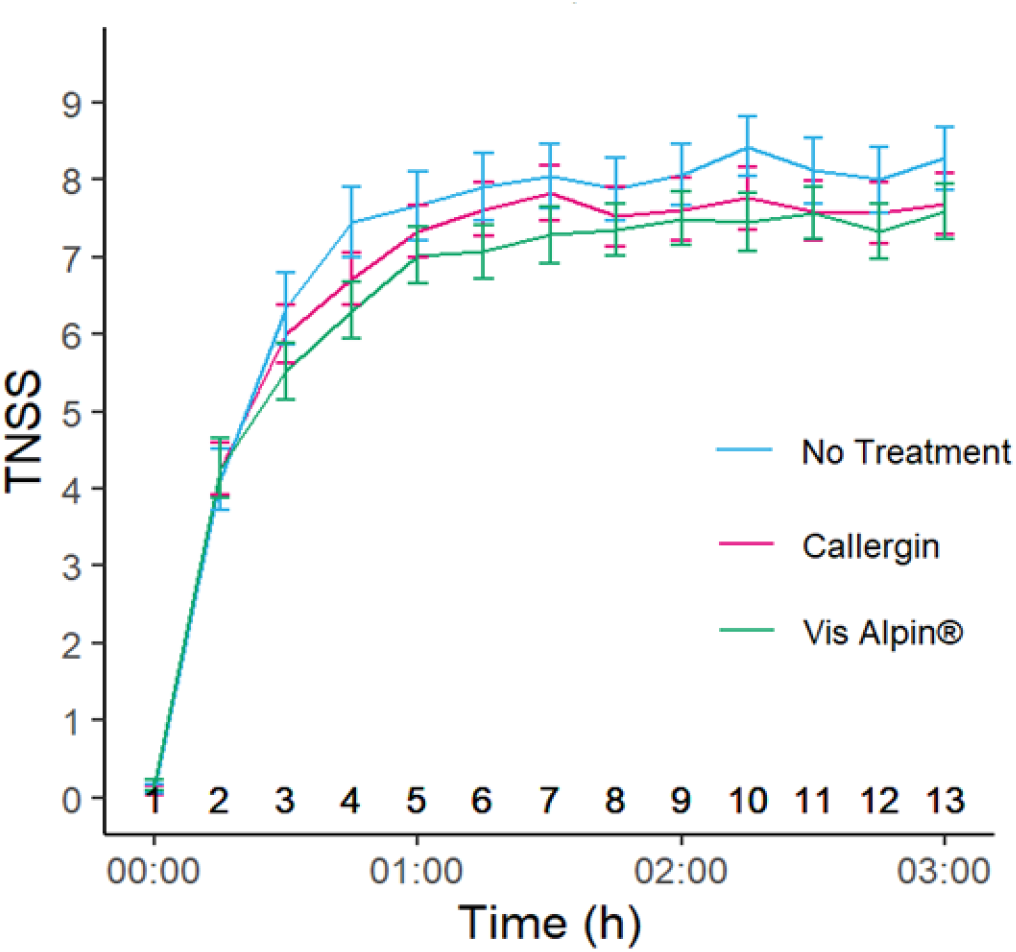
Average time course of TNSS following allergen exposure for the three treatment periods. Error bars indicate SEM.

As an objective parameter the weight of nasal secretions was evaluated and revealed a 13% reduction with Callergin treatment compared to no treatment (mean within-participant changes (g): untreated 2.85±2.63; Callergin 2.48±2.14 (Δ=0.37; 95%CI -0.10 to 0.84; p=0.119), None of the secondary endpoint analyses (TOSS, TRSS and nasal airflow) revealed any treatment differences in allergy symptoms (data not shown).

Post-hoc analysis evaluating TNSS at the end of the pollen challenge reaffirmed the Callergin effect on the relief of allergy symptoms. At the end of the three-hour allergen challenge, subjects receiving Callergin nasal spray showed significantly lower TNSS than those receiving no treatment (Δ=0.60; p=0.028) (Table 2). All individual nasal symptoms contributed to the Callergin effect, but rhinorrhea (Δ=0.26; p=0.013) and nasal congestion (Δ=0.17; p=0.076) to a larger extent. Fifty percent (50%) of the Callergin-treated subjects showed a TNSS of 8 or less compared with 8.5 or less in the untreated group. The top 25% most affected subjects experienced a TNSS of 9 or more in the Callergin-treated group and 10 or more in the untreated group (Table 3). Subjects treated with VisAlpin did not show a reduction in TNSS when compared to those receiving no treatment at 3 hours after the onset of the allergen challenge (data not shown). Weight of nasal secretions was reduced by 15 % measured at 3 hours, but did not reach significance (Table 2).

**Table 2:** Mean TNSS, its individual nasal symptoms, and nasal secretion weight at 3 hours. Difference from “no treatment” control is given as mean treatment difference with associated 95% CIs and P values.

|  | No<br>Treatment<br>(mean±SD) | Callergin<br>(mean±SD) | Mean<br>difference | 95% CI | P-Value<br>of Wilcoxon<br>Test |
| --- | --- | --- | --- | --- | --- |
| TNSS | 8.286±2.635 | 7.690±2.561 | 0.595 | -0.097, 1.287 | 0.028 |
| Itchy nose | 2.071±0.838 | 2.000±0.883 | 0.071 | -0.239,0.382 | 0.520 |
| Nasal congestion | 2.452±0.705 | 2.286±0.673 | 0.167 | -0.014,0.348 | 0.076 |
| Rhinorrhea | 2.095±0.850 | 1.833±0.853 | 0.262 | 0.067,0.457 | 0.013 |
| Sneezing | 1.667±0.928 | 1.571±0.941 | 0.095 | -0.178,0.369 | 0.387 |
| Nasal secretion (g) | 2.535±3.020 | 2.144±2.062 | 0.391 | -0.139,0.920 | 0.218 |

**Table 3:** Population analysis of TNSS scores.

|  | Median |  | 25%-Percentile |  | 75%-Percentile |  |
| --- | --- | --- | --- | --- | --- | --- |
|  | No Treatment | Callergin | No Treatment | Callergin | No Treatment | Callergin |
| TNSS | 8.5 | 8.0 | 7.0 | 7.0 | 10.0 | 9.0 |
| Itchy nose | 2.0 | 2.0 | 2.0 | 2.0 | 3.0 | 3.0 |
| Nasal congestion | 3.0 | 2.0 | 2.0 | 2.0 | 3.0 | 3.0 |
| Rhinorrhea | 2.0 | 2.0 | 2.0 | 1.0 | 3.0 | 2.0 |
| Sneezing | 2.0 | 1.5 | 1.0 | 1.0 | 2.0 | 2.0 |

In the safety population, a total of 11 adverse events occurred in 6 participants during the trial. The only adverse event reported more than once was headache. Importantly, the incidence of adverse events was comparable among treatment groups (Table 4). None of these events was serious or led to treatment withdrawal. Moreover, laboratory blood results did not reveal any clinically significant abnormalities.

**Table 4:** Safety profile. All adverse events started during a washout phase.

|  | No Treatment |  | Callergin |  | VisAlpin |  | All (n=47) |  |
| --- | --- | --- | --- | --- | --- | --- | --- | --- |
|  | No.(%) subjects with events | No. events | No.(%) subjects with events | No. events | No.(%) subjects with events | No. events | No.(%) subjects with events | No. events |
| Any adverse event | 3(6%) | 6 | 2(4%) | 2 | 2(4%) | 3 | 6 (13%) | 11 |
| Any treatment-related adverse event | 0 |  | 0 |  | 0 |  | 0 |  |
| Any serious adverse event | 0 |  | 0 |  | 0 |  | 0 |  |
| Any adverse event leading to withdrawal | 0 |  | 0 |  | 0 |  | 0 |  |

## Discussion

In this study, we evaluated the clinical performance of Callergin nasal spray in reducing allergic rhinitis symptoms in subjects with grass pollen allergy when compared to a “no-treatment” control. Our results show a trend towards decreased nasal symptoms and nasal secretion during the whole challenge period following a single application of Callergin nasal spray. Although the effect did not reach statistical significance in the prespecified analysis, it did so in a post-hoc analysis evaluating nasal symptoms at the end of the challenge period. There were no adverse reactions in subjects in any of the treatment groups.

Single prophylactic treatment with Callergin reduced patient reported allergic nasal symptoms by 0.6 symptom points at 3 hours. In a clinical study investigating the pharmaceutical effect of different treatments on patient reported allergic nasal symptoms (TNSS 0-12 point scale), Gross et al. defined a threshold of 0.23 units as minimum clinical important difference. ^9^ In the light of this threshold a TNSS reduction by 0.6 units, achieved with a single application of Callergin, is a clinically relevant improvement.

In terms of population percentiles, 50% of the patients treated with Callergin showed a TNSS of 8 or less whereas untreated subjects revealed a TNSS of 8.5 or less. Moreover, the top 25% most affected subjects experienced a TNSS of 9 or more in the Callergin group compared to 10 or more in the untreated group. These data indicate that most of the allergic patients had a benefit of prophylactic Callergin treatment.

Consistently, the weight of nasal secretions was reduced by 13% over 3 hours in Callergin-treated individuals. Nasal obstruction, a typical late reaction that takes place up to 6 hours after exposure ^4^, showed no difference across treatment groups when assessed by rhinomanometry. The Callergin effect, as evidenced in nasal symptoms, was not observed in eye or respiratory symptoms, which is unsurprising considering the product is a medical device acting on the local mucosal surface.

VisAlpin (a saline nasal spray) was chosen as a comparator because it is indicated as an add-on treatment for stuffy nose due to a cold or allergy. In a meta-analysis study, saline nasal irrigation produced a 28% improvement in nasal symptoms and a 62% reduction in medicine consumption in AR patients when performed daily over a period of up to 7 weeks.^10^ In our study, VisAlpin improved nasal symptoms somewhat, but not significantly. VisAlpin flushes out allergens from the nasal mucosa, while Callergin forms a long-lasting protective barrier on the nasal mucosa and thus is more appropriate as a prophylactic treatment.

Other barrier-forming nasal sprays that prevent allergen contact are currently on the market or under development for the treatment of AR. Tested substances include cellulose derivatives ^11^, clay mineral bentonite ^12^, thixotropic gel ^13^, petrolatum-based ointment ^14^, and lipid-based ointment.^15^ Of these, a hydroxypropyl methylcellulose powder (Nasaleze^®^), on sale since 1994, has been backed up by over 20 clinical studies. In a study conducted in a natural setting - during the pollen season - patients were asked to apply nasal puffs and document their symptoms daily. A 4-week therapy with Nasaleze^®^ reduced TNSS by 26% compared to placebo. ^16^ Other researchers evaluated the effect of a single application of Nasaleze^®^ on TNSS using two different experimental allergen challenges: either by direct instillation of an allergen into the nasal cavities or through an environmental challenge chamber (such as the one used in our study).

Results showed that a single application of Nasaleze^®^ before an intranasal dust mite challenge reduced TNSS by 41% over 4.5 hours in comparison to placebo.^17^ In contrast, Nehrig and colleagues found that a single application of Nasaleze^®^ before a 4-h grass exposure to pollen in an environmental challenge chamber reduced TNSS by 12% over 4 hours in comparison to a “no treatment” control.^12^ This difference in effect size suggests that the continuous allergen challenge is much harsher than the single intranasal allergen challenge normally used for investigating antiallergic nasal sprays. In fact, the extent of TNSS reduction by a single application of Nasaleze^®^ following a continuous allergen challenge is similar to that observed in our study with Callergin. Therefore, a more pronounced effect, similar to the one seen after multiple applications of Nasaleze (26% reduction in TNSS), can be expected with Callergin when it is used regularly during the pollen season. As Callergins’ main ingredient, iota-carrageenan, has an excellent safety profile and a long history of intranasal long-term applications also in the sensitive population (children, pregnant women, elderly), Callergin can be used regularly without any safety concerns, The long lasting barrier develops its protective effect immediately after application and lasts up to 3 hours, as shown by an immediate reduction of allergy symptoms which reaches significance at the end of the allergen challenge. As Callergin acts only locally without any pharmacological effect, the nasal spray can be used by vulnerable populations.

Strengths of this study include a crossover design, in which each patient acts as her/his own control, the random assignment to minimize possible effects from the order of treatment, and a zero dropout rate. The 7-day washout period was considered sufficient to eliminate any effects of previous exposure to the allergen, as TNSS returned to baseline values at the beginning of each treatment period. A possible limitation of this study is the comparison of Callergin to VisAlpin and a “no treatment” group, which does not allow complete blinding. However, the use of the blinded comparator allows for unbiased interpretation and confirms the findings of this study.

## Conclusion

In conclusion, Callergin nasal spray can be considered safe and effective in the relief of nasal symptoms in adults with grass pollen allergy.

## Data Availability

All data produced in the present study are available upon reasonable request to the authors

## Ethics Statement

The study was conducted in accordance with the ethical principles set forth in the Declaration of Helsinki and approved by the Ethics Committee of the City of Vienna (protocol code CAL_19_01, EK 19-276-1219).

## Acknowledgments

This research was sponsored by Marinomed. Medical writing and editorial support were provided by Joana Enes (Gouya Insights).

